# Implementing portable, real-time 16S rRNA sequencing in the healthcare sector enhances antimicrobial stewardship

**DOI:** 10.1101/2024.09.23.24314079

**Authors:** David Carlisle, Edward Cunningham-Oakes, June Booth, Andrew Frankland, Michael McDowell, Jack Pilgrim, Aleksandra Rzeszutek, Ceri Evans, Susan Larkin, Ang Li, Christopher Loftus, Merna Samuel, Nicola Scott, Luke Swithenbank, Victoria Owen, Alistair C Darby, Anna Smielewska

## Abstract

**Background:** Antimicrobial resistance (AMR) poses a significant global health challenge, resulting in over 1.27 million deaths in 2019 and projected to cause up to 10 million deaths annually in the future. To address this issue, the healthcare sector requires rapid and accurate bacterial identification, which is currently not readily available for effective antimicrobial stewardship. In a UK national first, we implemented 16S ribosomal RNA (rRNA) sequencing using Oxford Nanopore Technology (ONT) in an NHS setting to enhance diagnostic capabilities, aiming to reduce antibiotic misuse and improve patient outcomes.

**Methods:** We implemented 16S rRNA sequencing via ONT, running samples from seven NHS hospitals across Cheshire and Merseyside. We focused on samples from sterile sites, such as “pus”, “fluid”, and “tissue”, typically collected from critical care units. The assay was validated against traditional methods including Sanger sequencing and MALDI-TOF, with a turnaround time of 24-72 hours. Clinical impact was measured by analysing changes in antibiotic regimens and patient outcomes based on 16S assay results over a period of several months post-launch.

**Findings:** ONT 16S rRNA sequencing significantly impacted antibiotic treatment in 34.2% of cases, reducing patient stays and outperforming traditional methods by detecting additional bacterial organisms and identifying bacteria missed by reference labs. It provided species-level identification and confirmed non-infectious conditions in 5.4% of cases, aiding alternative treatment decisions. Its speed, cost-effectiveness, and minimal training requirements contributed to its successful integration into clinical practice.

**Interpretation:** The integration of ONT 16S sequencing into routine NHS diagnostics has significantly improved antimicrobial stewardship by offering a faster, more sensitive, and accurate bacterial identification method. Earlier use of this assay in cases where routine cultures are likely to fail could enhance patient outcomes further by enabling timely, targeted antibiotic therapies, reducing hospital stays, and curbing unnecessary antibiotic use.

## INTRODUCTION

Antimicrobial resistance (AMR) is one of the most critical health challenges of the 21st century^1^. This silent pandemic, facilitated by a stagnant antibiotic pipeline and the need for improved antimicrobial stewardship^2^, is already responsible for significant illness and death globally. Recent data indicate that in 2019 alone, 1.27 million deaths were linked to AMR, and this number is projected to reach up to ten million in the coming years^3^. AMR is also listed as one of the four chronic risks on the UK National Risk Register along with Climate Change, Risks posed by AI and Organised Crime^4^.

The same document recognises the key importance of both the reduction as well as the optimisation of the use of antimicrobials. Rapid and precise diagnostics are the first step towards these goals^5^. However, the current methods in routine diagnostic laboratories do not always facilitate this need. Traditional culture methods are unsuitable for fastidious or slow-growing organisms^6^ and samples post-antibiotic therapy, whilst molecular methods are restricted to a specific set of organisms^7^. Real-time, accredited clinical metagenomic assays that can be deployed locally and show real clinical impact are more important than ever^8^, given the increasing threat posed by antimicrobial resistance.

The 16S ribosomal RNA (rRNA) gene is ubiquitous in prokaryotes^9^. Direct and rapid diagnostics from clinical samples using 16S sequencing (metataxanomics) falls broadly under the umbrella of clinical metagenomics^8^. This method of rapid pathogen identification offers advantages for preventing antimicrobial resistance by enabling rapid pathogen identification. This allows tailoring of treatment and facilitates antimicrobial stewardship. Moreover, this approach is not limited by bacterial culture^10^ and is suitable for detecting complex polymicrobial infections, including rare organisms in patients experiencing polypharmacy^11^.

Here, we outline the implementation of whole gene 16S sequencing using Oxford Nanopore Technology with fast turnaround time (24-72 hours) and lean bacterial diagnosis and identification for improved antimicrobial stewardship. We describe the impact of this technology on inpatient time and antibiotic course of treatment across seven different hospitals and provide extensive benchmarking against externally accredited standards. This work will serve as a guide to those seeking to implement pathogen diagnostics as clinically-accredited assays, thereby bridging the gap between genomics and clinical diagnostics.

## METHODS

### Sample collection for assay validation

A total of 109 samples were included in the assay validation part of the project. Of these, 104 were surplus clinical samples (Table 1) and five were from an external quality control scheme; Quality Control for Molecular Diagnostics (QCMD)^12^. All clinical samples used were surplus material that had been stored at -20°C by the routine diagnostic Microbiology laboratories involved in the project.

**Table 1:**
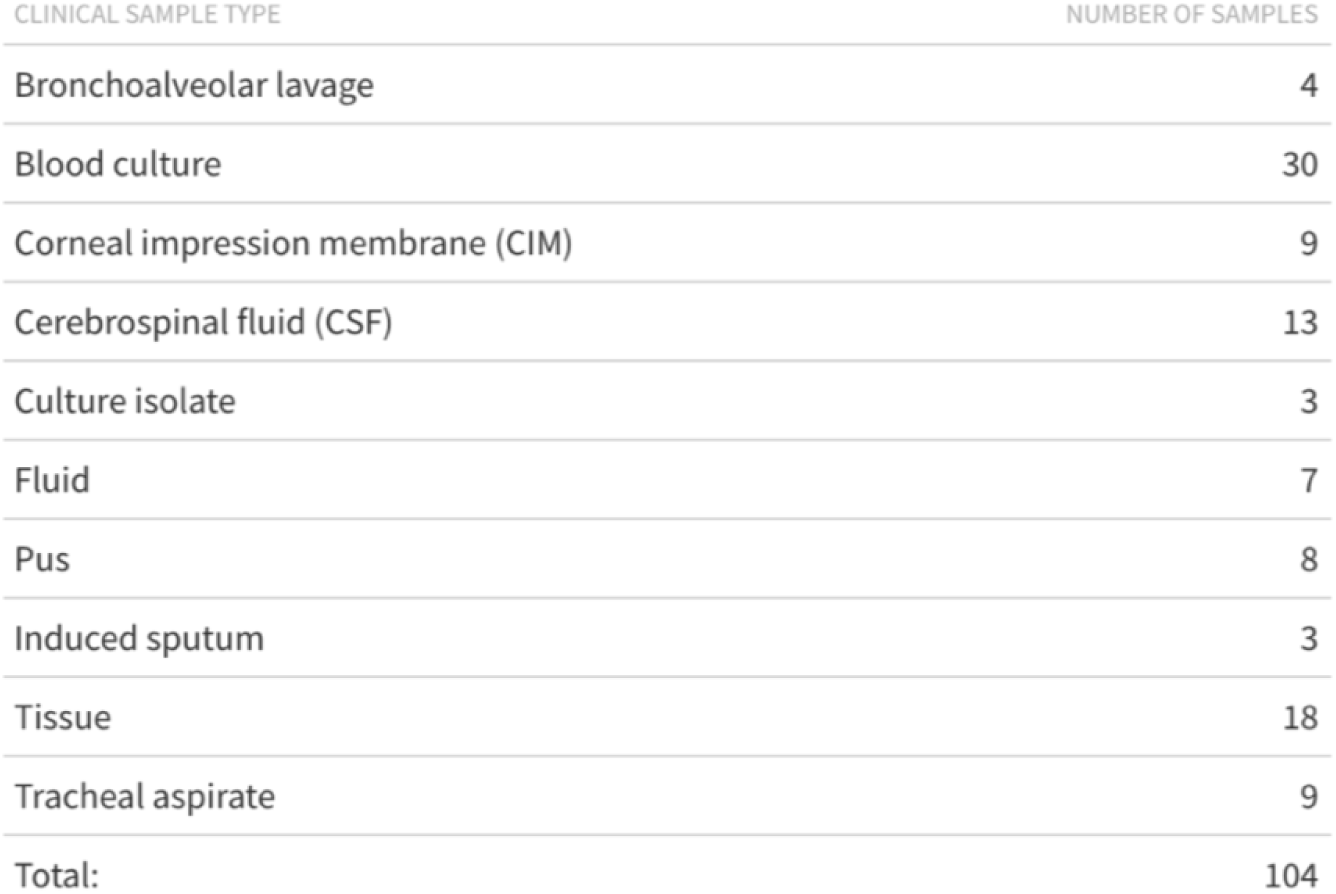
Summary of clinical samples and their site of origin.

| CLINICAL SAMPLE TYPE | NUMBER OF SAMPLES |
| --- | --- |
| Bronchoalveolar lavage | 4 |
| Blood culture | 30 |
| Corneal impression membrane (CIM) | 9 |
| Cerebrospinal fluid (CSF) | 13 |
| Culture isolate | 3 |
| Fluid | 7 |
| Pus | 8 |
| Induced sputum | 3 |
| Tissue | 18 |
| Tracheal aspirate | 9 |
| Total: | 104 |

### Read-based 16S microbiome profiling and consensus 16S pathogen sequence generation

We developed a simple bioinformatic workflow for analysis of 16S data both a read-by-read basis (for both low, and high-abundance samples), and consensus fasta files (high-abundance samples). The workflow was developed to account for the inherent variability in the bioburden of clinical samples, while also taking into account varying computational infrastructures across different Trusts and limited bioinformatics training.

Additional confidence in the validity of results for low-abundance samples was provided by clinical interpretation of results in relation to patient outcome, and additional diagnostic data where available.

Nanopore sequencing data (reads) were obtained from the experiment in FASTQ format. NanoFilt (v2.8.0) was then used to quality-filter reads based on the specified quality thresholds (Q12, minimum 850bp, maximum 1500bp). This threshold was selected based on the recommended input for consensus fasta generation using amplicon_sorter.py (version = ‘2023-06-19’). Reads not meeting the quality threshold were excluded from further analysis, and the remaining reads were merged.

Kraken2 (v2.0.7-beta) was then used to classify the selected Nanopore reads taxonomically, using Greengenes Database Release 13_5, with a confidence threshold of 0.1. These results were visualised in the Pavian web browser (https://fbreitwieser.shinyapps.io/pavian/), to enable accessible interpretation, and comparison of clinical samples and controls.

Amplicon sorter (v2022-03-28) was then used to group reads from samples based on their gene associations. Samples meeting the following criteria were then used to generate consensus fasta files: 850-1500bp length read and 1000 reads. Consensus fasta files were then classified using rRNA/ITS database via NCBI BLAST. Results with a minimum of >95% percentage identity and >90% coverage in comparison to a database entry were then taken forward for identification according to the The Clinical & Laboratory Standards Institute (CLSI) guideline MM-18A (Vol. 28 No 12): Interpretive Criteria for Identification of Bacteria and Fungi by DNA Target Sequencing.

### Cross-comparison of sequencing to gold-standard clinical results

Our workflow underwent External Quality Assessment (EQA) with Quality Control for Molecular Diagnostics (QCMD). QCMD provided a series of bacterial samples from three different distribution cycles (2016, 2017 and 2021), for which the correct bacterial identification was known. DNA was extracted from these samples, alongside all clinical samples (described in Table 1) and assessed using a previously accredited method for lean bacterial diagnosis (Sanger sequencing). 27F and 797R primers were used to amplify a ∼770 bp region of the 16S gene. Amplicon integrity and size was then confirmed via 1% agarose gel electrophoresis. Products were subsequently purified with Exo-SAP IT (Life Technologies, Waltham, MA, USA) by adding 5 µL of the enzyme to the PCR product, followed by incubation at 37°C for 15 minutes and 80°C for another 15 minutes.

Subsequent PCR was conducted using 1 µL of the purified product with the BigDye™ Terminator v1.1 Cycle Sequencing Kit to incorporate fluorescently tagged bases^[12]^. Post-PCR, amplicons were purified again using a two-step ethanol precipitation process: first with 24 µL absolute ethanol and 1 µL 3M NaAc, followed by centrifugation at 2000 x g for 20 minutes, and then with 75 µL 70% ethanol and centrifugation at 2000 x g for five minutes. The precipitated DNA was resuspended in 10 µL Hi-Di Formamide (Life Technologies) and sequenced on an ABI 3500 genetic analyser (Life Technologies).

Consensus sequences were generated using SeqScape™ Software (Life Technologies) and submitted to BLAST (NCBI, Bethesda, MD, USA) to identify organisms at the genus or species level, following CLSI guidelines. Results were stored for later comparison. If sequencing did not yield a result, samples were cultured on Columbia blood agar (Merck & Co., Rahway, NJ, USA) and identified using the Bruker Microflex® LRF MALDI-TOF benchtop analyser. Results for 2023 from both ONT and Sanger sequencing were then reported to QCMD.

### Determining the clinical demand and impact of introducing ONT diagnostics

Data was collected from 7 different hospitals including a paediatric and a cancer hospital from the introduction of the test on 01/05/23 to 15/02/24.

For each case, the following data were obtained from the laboratory database and patient notes: date, source and result of sample; basic patient demographics (including sex and date), date of patient admission, discharge and location when sample was taken. Additional data collected from patient notes included the clinical impact of the test, specifically focusing on no change in patient treatment: no change/confirmation of current antimicrobial regimen/confirmation of non-infectious cause vs change in antimicrobial treatment such as stop/start or narrow antibiotic therapy.

For inpatients, linear regression was subsequently used to determine if there was a relationship between the length of time (days) from patient admission to 16S result and the overall length of patient stay depending on the clinical impact of the result i.e. change or no change in treatment (control).

## RESULTS

### Assay validation

#### 16S sequencing using ONT improves results for accreditation from the previous clinical pathway, and accurately characterises polymicrobial clinical samples

All five samples produced results in line with those issued by QCMD, including an additional educational sample provided by QMCD that had two different organisms present (*Klebsiella pneumoniae* & *Acinetobacter baumannii*). This sample, when originally tested and reported in 2021 via the previous method, was reported as ‘unable to sequence’ due to the mixed composition of the sample, however when tested as part of this exercise, the two different organisms were both correctly identified. These results are summarised in Table 2.

**Table 2:**
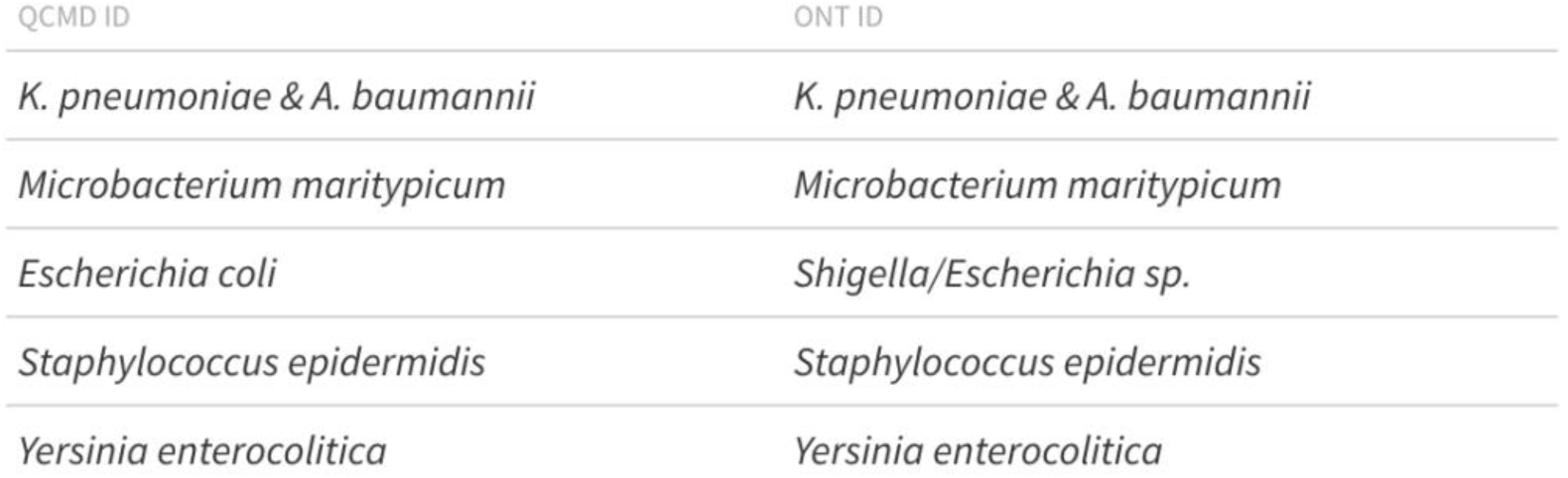
Comparison of taxonomy expected for accreditation and ONT results.

The ONT protocol produced organism matches or improved diagnosis in all clinical samples included in assay validation. For the 8 samples where the previous Sanger sequencing method failed to produce a sequence, culture and MALDI-TOF were used to obtain an organism for comparison, with all 8 identifications matching. Among the 24 samples reported as negative by the reference laboratory, the ONT assay results were consistent for 21 samples, but bacterial DNA was amplified for 3 samples, including a *Fusobacterium sp*. in a CSF sample and *Pseudomonas aeruginosa* in two fluid samples.

In samples referred to the reference laboratory, targeted PCR was used instead of the 16S sanger sequencing. In these cases ONT could be used to identify the reported organisms on the basis of 16S alone, and identified three additional organisms (Table 3).

**Table 3:**
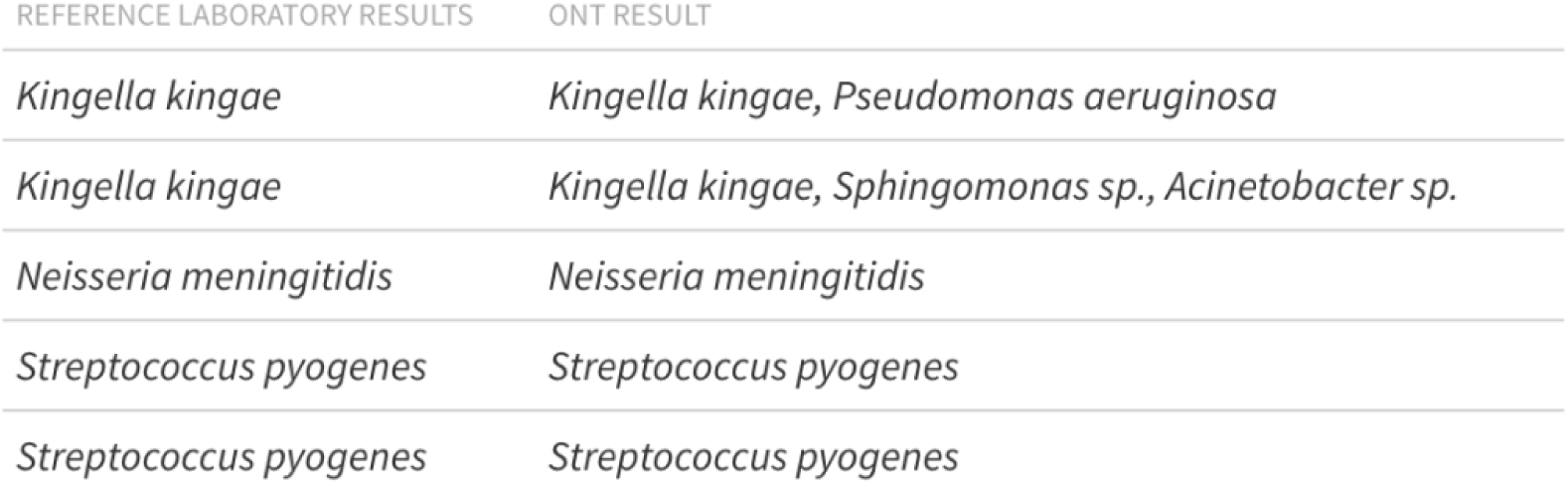
Comparison of reference laboratory and ONT results.

### Patient and sample demographics and assay impact

#### The largest demographic for sample requests were inpatients and patients aged 50-69

Patients originated from 7 different trusts, including acute trusts and specialist neurological, surgical, oncology, and paediatric centres (210 samples; see Table 4 and Figure 1). In terms of age demographics, the majority of patients were in the 50-69 age group (Males = 21.2% of patients, Females = 18.2% of patients). The smallest demographic sampled was in the 17-25 age group, (Males = 1.3% of patients, Females = 3.0% of patients). In five out of seven age groups, more patients were male than female.

**Figure 1:**
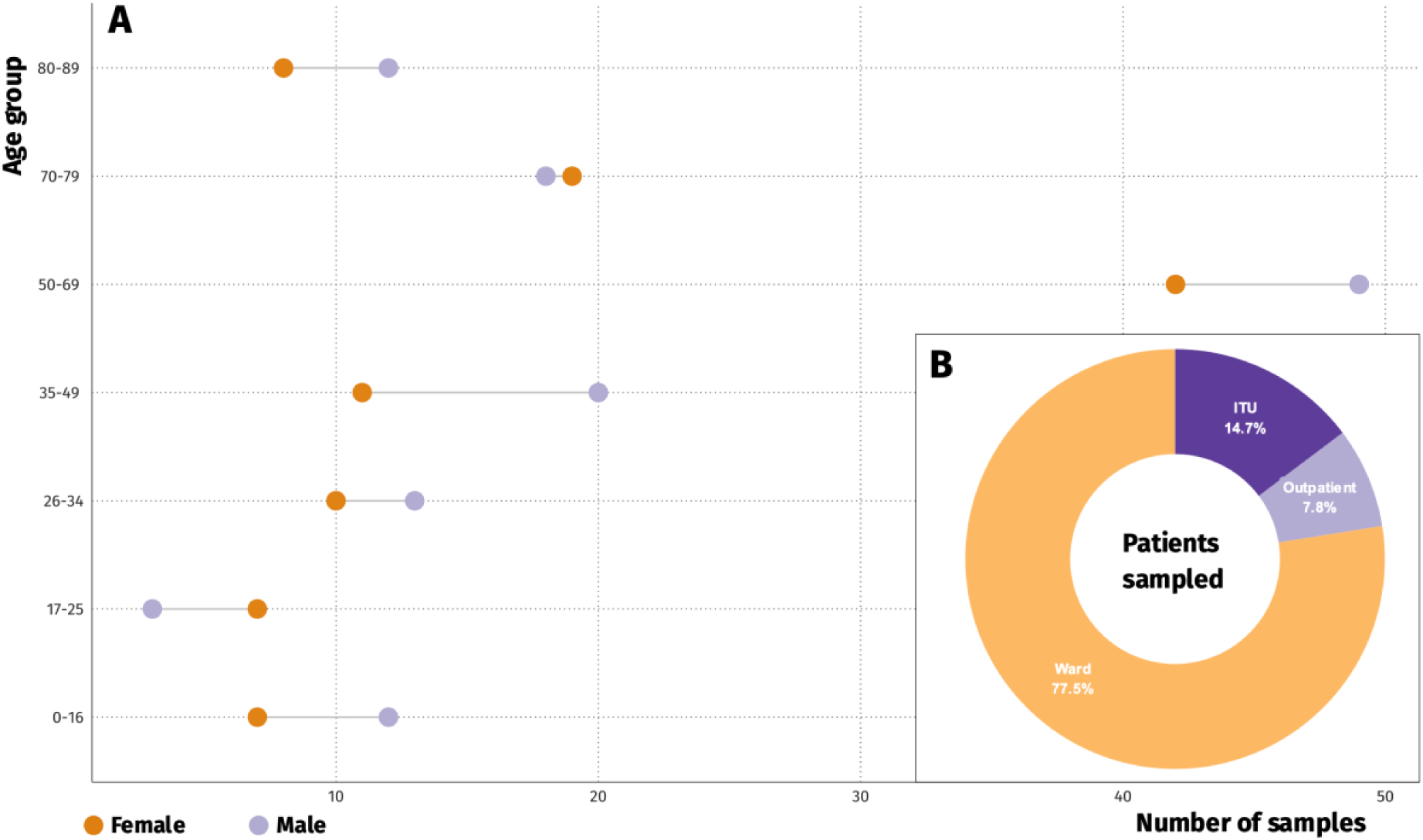
(A) Sex and age distribution of (210) patients. Patients originated from 7 different trusts including acute trusts and specialist neurological, surgical, oncology and paediatric centres. (B) Pie chart summarising patient locations when the sample was taken. The majority of outpatient requests originated from Ophthalmology. 14.7 % of samples originated from ITU, for comparison the largest acute trust (LUHFT) included in the data collection ∼ 5.5% critical care beds, the sampling is clearly towards critical care admissions, reflecting sicker patients with longer stays.

**Figure 2:**
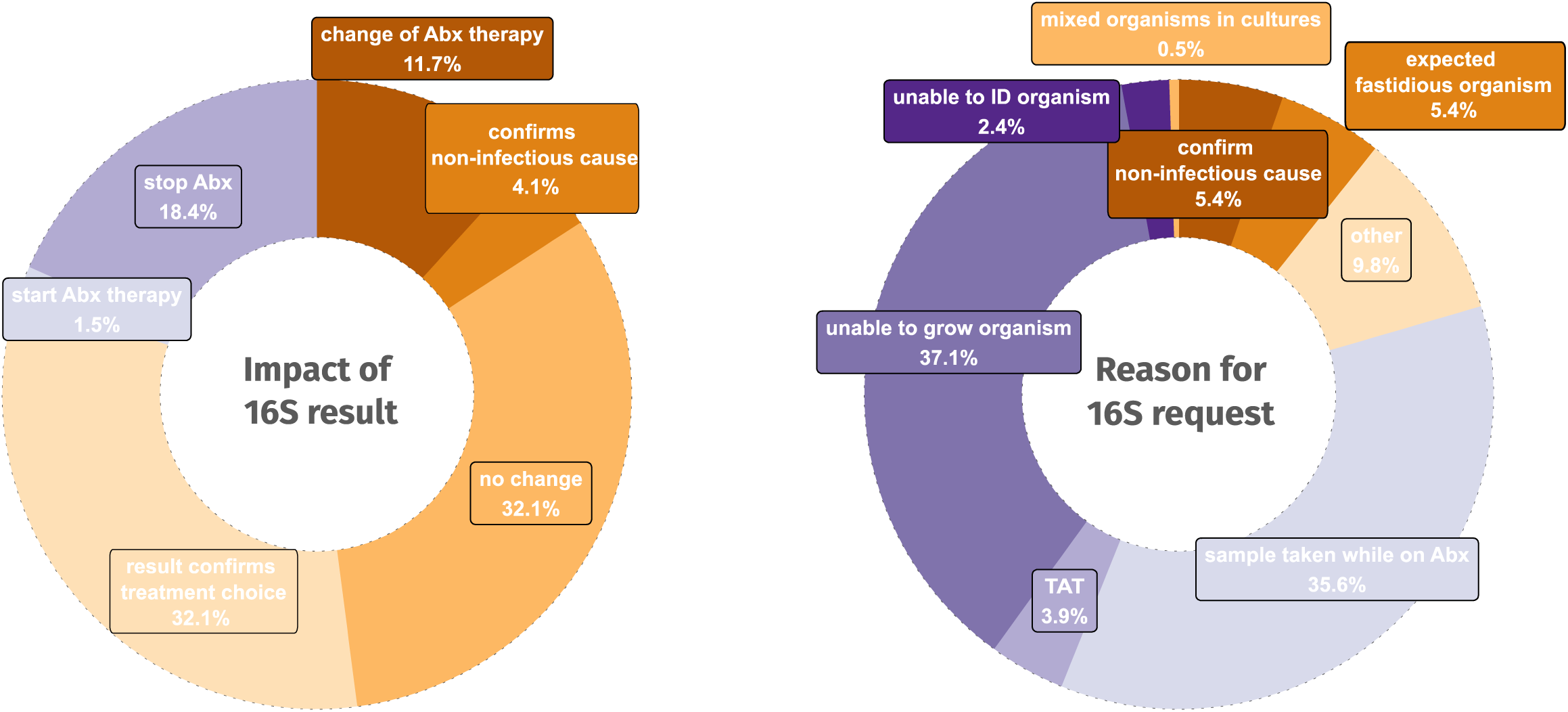
Requests for 16S analysis support antimicrobial stewardship efforts and are most common when bacterial culture is unsuccessful. (A) Summary of clinical decision-making post 16S result. The majority of outcomes either confirmed treatment choice or made no difference, however the test has resulted in a change in therapy in 35.7% of cases and in stopping or narrowing the antibiotic regimen in 34.2% of cases. (B) Summary of the 16S requests. The majority of requests originated from failure to grow with conventional culture methods.

**Figure 3:**
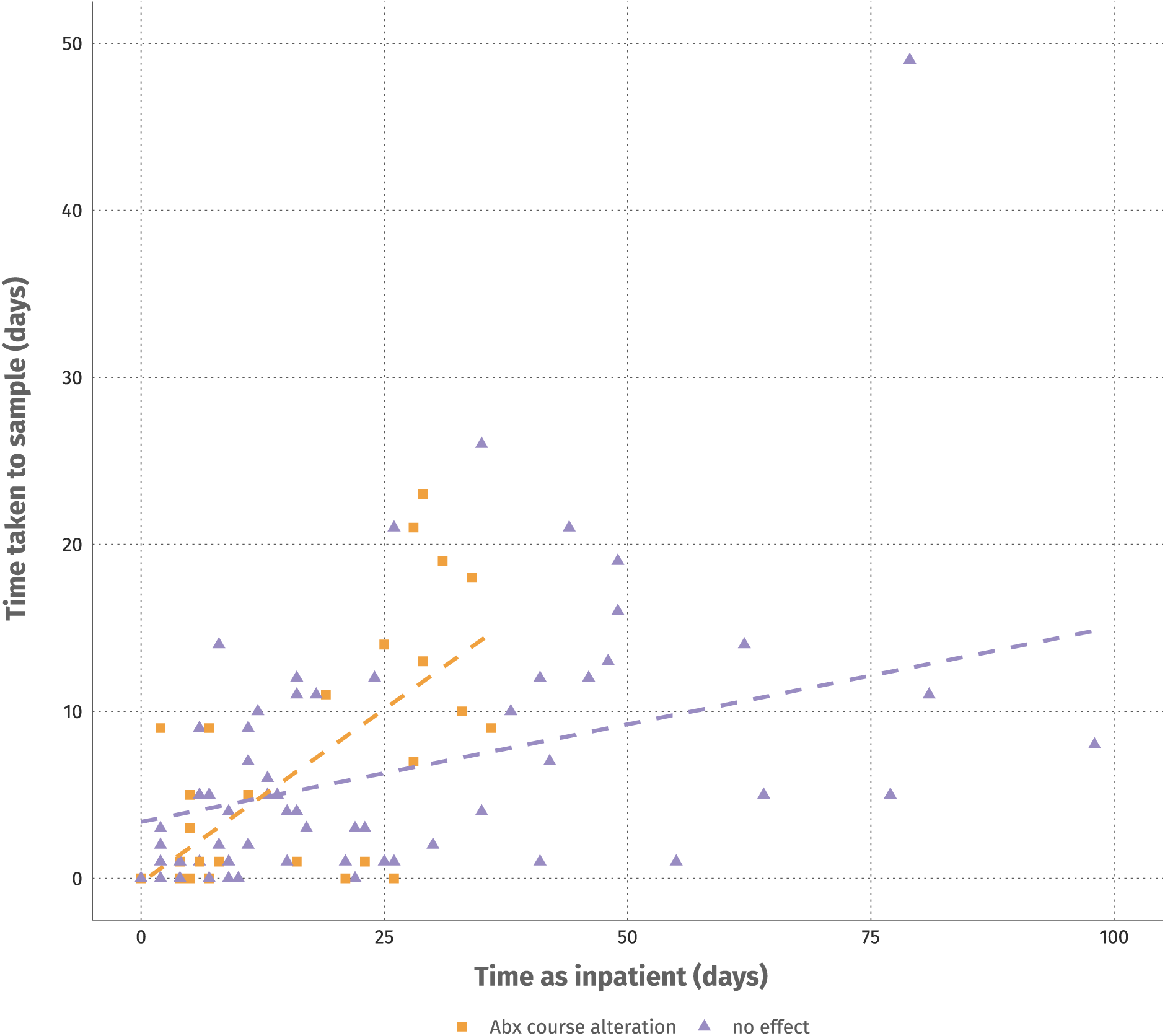
Faster 16S sampling requests decrease inpatient time and antibiotic exposure. Linear regression showing the correlation between time taken to sample and time as an inpatient in cases where 16S sampling resulted in altered antibiotic treatment (r^2^ = 0.4) or where sampling did not affect treatment (r^2^ = 0.03).

**Table 4:** Demographics of patients sampled for 16S.

| AGE GROUP | TOTAL | MALES (N) | MALES (%) | FEMALES (N) | FEMALES (%) |
| --- | --- | --- | --- | --- | --- |
| 0-16 | 19 | 12 | 5.2 | 7 | 3 |
| 17-25 | 10 | 3 | 1.3 | 7 | 3 |
| 26-34 | 23 | 13 | 5.6 | 10 | 4.3 |
| 35-49 | 31 | 20 | 8.7 | 11 | 4.8 |
| 50-69 | 91 | 49 | 21.2 | 42 | 18.2 |
| 70-79 | 37 | 18 | 7.8 | 19 | 8.2 |
| 80-89 | 20 | 12 | 5.2 | 8 | 3.5 |

The majority (77.5%) of samples were from patients in wards, followed by intensive care units (ITUs; 14.7%) and outpatient clinics (7.8%). The majority of outpatient requests originated from Ophthalmology. The largest acute trust (LUHFT) included for data collection has ∼5.5% critical care beds. As such, the number of samples referred from ITUs reflects a focus on critical care admissions, including sicker patients with longer stays.

#### Faster 16S sampling requests decrease inpatient time and antibiotic exposure

The majority of outcomes post 16S result either “confirmed treatment choice” or “made no difference”, however the test has resulted in a change in therapy in 35.7% of cases and in stopping or narrowing the antibiotic regimen in 34.2% of cases. The adjusted R² for altered therapy post 16S result was 0.4, indicating a positive correlation, the strongest positive correlation was observed between rationalisation of antibiotic usage and length of stay, with an R² of 0.56, meaning 56% of the variability could be accounted for by the delay taken to request a 16S sample. This suggests that longer times to request a 16S sample may be associated with longer hospital stays in this group (34.2% of patients sampled). As expected, there was no significant correlation between length of stay in cases where the outcome had no effect, with an R² of 0.03. Although confirming a non-infectious cause did not impact the length of patient stay, it remains an important factor in antibiotic stewardship as well as for subsequent patient treatment. These findings highlight the potential impact of timely 16S sample requests on reducing hospital stays, particularly when antibiotics are changed.

## DISCUSSION

In a national first, we have successfully implemented 16S sequencing using Oxford Nanopore Technology (ONT) as part of an NHS-accredited assay for rapid bacterial diagnosis and identification. This method offers a turnaround time (TAT) of 24-72 hours, which is significantly faster than traditional methods (e.g. 16S PCR from culture, or MALDI-TOF for bacterial identification) or the previous method that took approximately seven days from sample receipt. Critically we have been able to show that the assay results had influenced clinical decision-making, leading to changes in antibiotic regimens in 34.2% of cases with a potential to influence the length of patient stays. The reduced training time and staffing requirements coupled with the associated cost-effectiveness of this method were also notable.

ONT offers several advantages over traditional Sanger sequencing, allowing the sequencing of the entire 16S gene in a single contig and eliminating the need for sequence reassembly^13^. Here, we have shown that we were able to enhance service delivery without sacrificing reliability, which is a common concern for culture-independent assays^14^. The only discrepancies reported during assay validation and verification were cases where ONT detected additional organisms compared to previous methods or provided species-level identification where the older methods could only offer genus-level classification. The enhanced sensitivity during method validation resulted in four instances where the ONT method detected additional bacterial organisms compared to the previous method and three cases where ONT identified bacteria in samples that the reference laboratory had reported as negative. These findings had the potential to pose challenges for clinicians, as the presence of multiple microorganisms could complicate interpretation^15^, which is often the case with metagenomic approaches^16^. These results were subsequently used in an informed education campaign for clinical teams before the launch of the assay, ensuring safe delivery of results.

With the launch of the 16S ONT assay, we have successfully demonstrated the feasibility of integrating clinical metagenomics into routine diagnostics. Metagenomic approaches tend to be novel methods that are routinely confined to very complex cases with usage guidelines determined by lack of accessibility, prohibitive costs and long turnaround times^8^. Our aim was to develop an accessible metataxonomic assay, free of these constraints, while collecting data on the usage of the assay as well as on the subsequent impact on patient care with a view of introducing evidence-based guidelines optimising the use of these highly effective tools.

To this end, we have collected data both of sample and patient demographics throughout the first 9 months since the assay launch, as well as on the clinical impact as determined by the ward teams of the results of the 16S assay. Since its introduction, the assay has been adopted by seven hospitals across Cheshire and Merseyside, with a noticeable bias towards requests from critical care units, possibly reflecting the complexity of cases requiring longer hospital stays and more intricate antibiotic regimens. Samples from all sterile sites were accepted for 16S with “pus”, “fluid” and “tissue” accounting for the majority of these. These reflect infectious conditions that would both be challenging for antibiotic penetration such as abscesses as well as potentially requiring prolonged antibiotic treatment such as in, for example, joint fluids and biopsies or infected grafts^17^.

Unsurprisingly, the primary reason for 16S testing was the failure of conventional culture methods to identify the organism, or a reduction of the likelihood that the culture would be successful, due to previous therapy. This points to a systematic delay in the request of the 16S assay, although with an inevitable sampling bias as the assay would be requested if routine methods failed to produce a result or an antibiotic regimen was deemed to be ineffective. However, we have also demonstrated that in just over a third of the cases, the longer time to request a 16S sample was associated with longer hospital stays. This conclusion coupled with the use of the 16S assay in more complex cases, and with harder to treat infection foci, points to a subset of patients where a more timely 16S request could have the most significant impact. It is also important to note the 5.4% of the cases, where a negative 16S assay was used to confirm a non-infectious cause, likely facilitating an alternative treatment, although not directly affecting time to discharge.

In conclusion, it is recognised that rapid and effective diagnostics are the first step towards improved antimicrobial stewardship leading to both a reduction and an optimisation of the use of antibiotics. To this end we have shown that the optimised ONT 16S workflow offers a sensitive, specific, and rapid method for identifying bacterial organisms. The workflow successfully addresses the limitations of the traditional Sanger method and has become an invaluable component of our routine diagnostic repertoire. Future applications of this assay are being explored, including further optimisation for different sample types, such as microbial keratitis and water contamination monitoring at local hospitals. We have additionally demonstrated that more research is required in the identification of samples where routine culture would be less successful, including high probability of fastidious organisms such as mycobacteria or previous therapy, taking into account both factors relating to the patient as well as the nature of the sample. This data would lead to identifying a group of patients that would benefit from a more timely metagenomic test with a positive correlation with patient hospital stays and better antibiotic stewardship.

### Ethical approval

This quality improvement project utilised surplus diagnostic materials and reference material provided for this purpose to improve service quality and as part of the diagnostic laboratory quality assessment. It is not classified as research under the UK Clinical Research Collaboration. Therefore, it does not require ethical approval. However, the work was approved by Liverpool Clinical Laboratories (LCL), Liverpool University Hospitals NHS Foundation Trust, and NHS England as part of UK Health Security Agency Standards for Microbiology Investigations (UK SMI) Evaluations, Verifications, and Validations of Diagnostic tests.

### Funding information

Liverpool Clinical Laboratories (LCL) funded this work. E.C.-O. and A.C.D. are affiliated to the National Institute for Health Research (NIHR) Health Protection Research Unit in Gastrointestinal Infections at University of Liverpool, in partnership with the UK Health Security Agency (UKHSA), in collaboration with University of Warwick. E.C.-O. and A.C.D. are based at The University of Liverpool. The views expressed are those of the author(s) and not necessarily those of the NIHR, the Department of Health and Social Care or the UK Health Security Agency.

## Data Availability

All data produced in the present study are available upon reasonable request to the authors.

